# Elevated Plasma Matrix Metalloproteinase-8 associates with Sputum Culture Positivity in Pulmonary Tuberculosis

**DOI:** 10.1101/2021.11.15.21265734

**Authors:** N.F. Walker, F. Karim, M.Y.S. Moosa, S. Moodley, M. Mazibuko, K. Khan, T.R. Sterling, Y.F. van der Heijden, A.D. Grant, P.T. Elkington, A. Pym, A. Leslie

## Abstract

Current methods for tuberculosis (TB) treatment monitoring are suboptimal. We evaluated plasma matrix metalloproteinase (MMP) and procollagen III N-terminal propeptide concentrations before and during TB treatment as biomarkers. Plasma MMP-1, -8 and -10 significantly decreased during treatment. Plasma MMP-8 was increased in sputum *Mycobacterium tuberculosis* culture positive relative to culture negative participants, prior to (median 4609 pg/ml, IQR 2353-9048 vs 775 pg/ml, IQR 551-4920, p=0.019) and after 6 months (median 3650, IQR 1214-3888 vs 720, IQR 551-1321, p=0.008) of TB treatment. Consequently, plasma MMP-8 is a potential biomarker to enhance TB treatment monitoring and screen for possible culture positivity.

## Background

Tuberculosis (TB) is a major cause of morbidity and mortality worldwide, causing an estimated 9.9 million cases and approximately 1.5 million deaths in 2020, disproportionately affecting low resource settings (1). The treatment success rate for people receiving first line TB treatment was only 86% in 2019. For people living with HIV and those with multidrug-resistant (MDR) /rifampicin-resistant TB, treatment success rates are considerably lower (77% and 59% respectively) (1).

Monitoring response to TB treatment is a challenge for national TB programmes, especially in resource-limited settings. World Health Organization (WHO) recommendations are for repeat sputum smear microscopy for acid-fast bacilli after two months of TB treatment for people with a new diagnosis of pulmonary TB on first line treatment. Sputum culture for *Mycobacterium tuberculosis* (*Mtb*) is reserved for cases where sputum smear positivity persists or develops/recurs later in treatment (2). Patients who are sputum smear positive at diagnosis but smear negative at month two are recommended to have repeat smears at the end of month five and six. Both sputum smear and culture require appropriate laboratory facilities and trained personnel. Culture is limited by considerable time to result availability. Poor specificity for viable organisms precludes use of molecular amplification techniques such as Xpert MTB/RIF for treatment monitoring (3). All monitoring reliant on sputum production has diminished utility in sputum non-productive patients, including those who are too unwell, those with extrapulmonary TB and those whose cough has resolved on treatment. The WHO End TB strategy highlighted the need for new tools, including non-sputum-based diagnostics, to support effective patient-centred care (4).

Matrix metalloproteinases (MMPs) are host enzymes collectively capable of degrading the lung extracellular matrix at neutral pH, with broad immunological functions. They are tightly regulated *in vivo*, but have the potential to cause immunopathology (5). We have previously demonstrated *in vitro*, using animal models and in observational human studies, that MMP dysregulation is a key feature of TB immunopathology (6-10). We have shown that sputum MMP-1 and MMP-8 are significantly elevated in TB patients at diagnosis compared to controls, irrespective of HIV serostatus (7-9). In plasma, MMP-1, MMP-8 and procollagen III N-terminal propeptide (PIIINP), a matrix degradation product released during collagen turnover, are elevated in TB patients compared to healthy or respiratory symptomatic controls (8, 11). In patients without HIV infection, elevated sputum MMP-1, -2, -3 and -8 decrease after just two weeks of TB treatment (12). Here, in an exploratory study, we evaluated plasma MMPs and PIIINP concentrations and their association with sputum smear and culture status in a cohort of South African TB patients longitudinally, to determine the potential for use as novel peripheral biomarkers of TB treatment response.

## Methods

This was a retrospective analysis of the Collection of Sputum, Urine and Blood Samples for Research (CUBS) Study. CUBS prospectively recruited participants in health facilities in eTheKwini municipality, KwaZulu-Natal, South Africa. ETheKwini is one of 11 districts in the province and has a high TB incidence and HIV prevalence. Eligibility for CUBS was determined by age 18 years or above and ability to give written informed consent. Inclusion in this analysis required a diagnosis of TB (clinical and/or microbiological) leading to TB treatment initiation. All CUBS participants enrolled at the Prince Cyril Zulu Communicable Disease Centre (December 2013 - May 2014) were included in this study. Plasma samples were collected at baseline (TB diagnosis, prior to treatment initiation) and study visits at the end of month 2 (week eight) and month 6 (week 24) following TB treatment initiation.

HIV testing was offered if HIV status was not known. Sputum (routine or induced) was collected for mycobacterial analysis including smear, *Mtb* culture and drug susceptibility testing (DST) at baseline, month 2 and month 6 visits. Culture was performed on solid (7H11) and liquid (MGIT) media. DST was performed for isoniazid, rifampicin, ethambutol, streptomycin, ofloxacin and kanamycin. Plasma was collected in EDTA tubes at each timepoint. Plasma MMP-1, -3, -8, -9, and -10 were quantified by Luminex array (Bio-Rad Bio-Plex 200, assay from R&D Systems, UK) and PIIINP by ELISA (Cloud clone corp, China). The study was approved by University of KwaZulu-Natal and London School of Hygiene & Tropical Medicine research ethics committees (REFs BE022/13 and 11710 - 1 respectively).

Analysis was performed in Prism 8 (GraphPad, USA). Comparisons between groups were by Mann-Whitney U or Fisher’s exact test. Diagnostic accuracy was assessed by receiver operating characteristic (ROC) curve analysis. Associations between analytes were assessed by Spearman correlation.

## Results

Participant (n=85) characteristics at TB diagnosis are reported in Table 1. HIV serostatus was known to be positive in 43.5% (n=37) and unknown in 11.8% (n=10). The majority of participants were male (72.9%, n=62). TB diagnosis was confirmed on sputum culture in 91.8% (n=78). Baseline DST results were available for 80% (n=68) participants and 86.8% (n=59) of these were fully sensitive.

**Table 1.** Participant demographic and clinical characteristics

|  |  |
| --- | --- |
| **Total participants, n** | **85** |
| Female n (%) | 23 (27.1) |
| Age, median years (IQR) | 35 (28.5-42.0) |
| HIV positive, n (%) | 37 (43.5) |
| HIV status unknown n (%) | 10 (11.8) |
| Smear positive at diagnosis, n (%) | 43 (50.6) |
| Culture positive* at diagnosis, n (%) | 78 (91.8) |
| *Mtb* resistance test results at diagnosis |  |
| Fully sensitive, n (%) | 59 (69.4) |
| Isoniazid mono-resistant, n (%) | 4 (4.7) |
| Rifampicin mono-resistant, n (%) | 2 (2.4) |
| Streptomycin mono-resistant, n (%) | 1 (1.2) |
| Rifampicin & isoniazid resistant (MDR), n (%) | 2 (2.4) |
| Drug susceptibility results not available, n (%) | 17 (20.0) |

\*defined by a positive sputum culture for *Mtb*, on liquid or solid media, or both

### Plasma collagenases & PIIINP decrease during TB treatment

Plasma MMP & PIIINP concentrations during TB treatment are reported in the Figure, panels A-G. The collagenases, MMP-1 (interstitial collagenase) and -8 (neutrophil collagenase) decreased significantly between baseline and month 2 visits, as did the stromelysin, MMP-10. Concordantly, the matrix degradation product PIIINP, which is released during collagen turnover, decreased over the first two months of TB treatment. No further significant reductions between month 2 and month 6 were observed. Conversely, plasma MMP-9 significantly increased between baseline and month 2, whilst plasma MMP-3 and MMP-7 did not change. Assessing correlations between analytes including data from all timepoints revealed a positive correlation between plasma PIIINP and the collagenases MMP-1 (r=0.759, p<0.001) and MMP-8 (r=0.224, p<0.001). MMP-1 and MMP-8 were also positively correlated (r=0.377, p<0.001). Full correlation results are reported in Supplementary Table S1.

**Figure:**
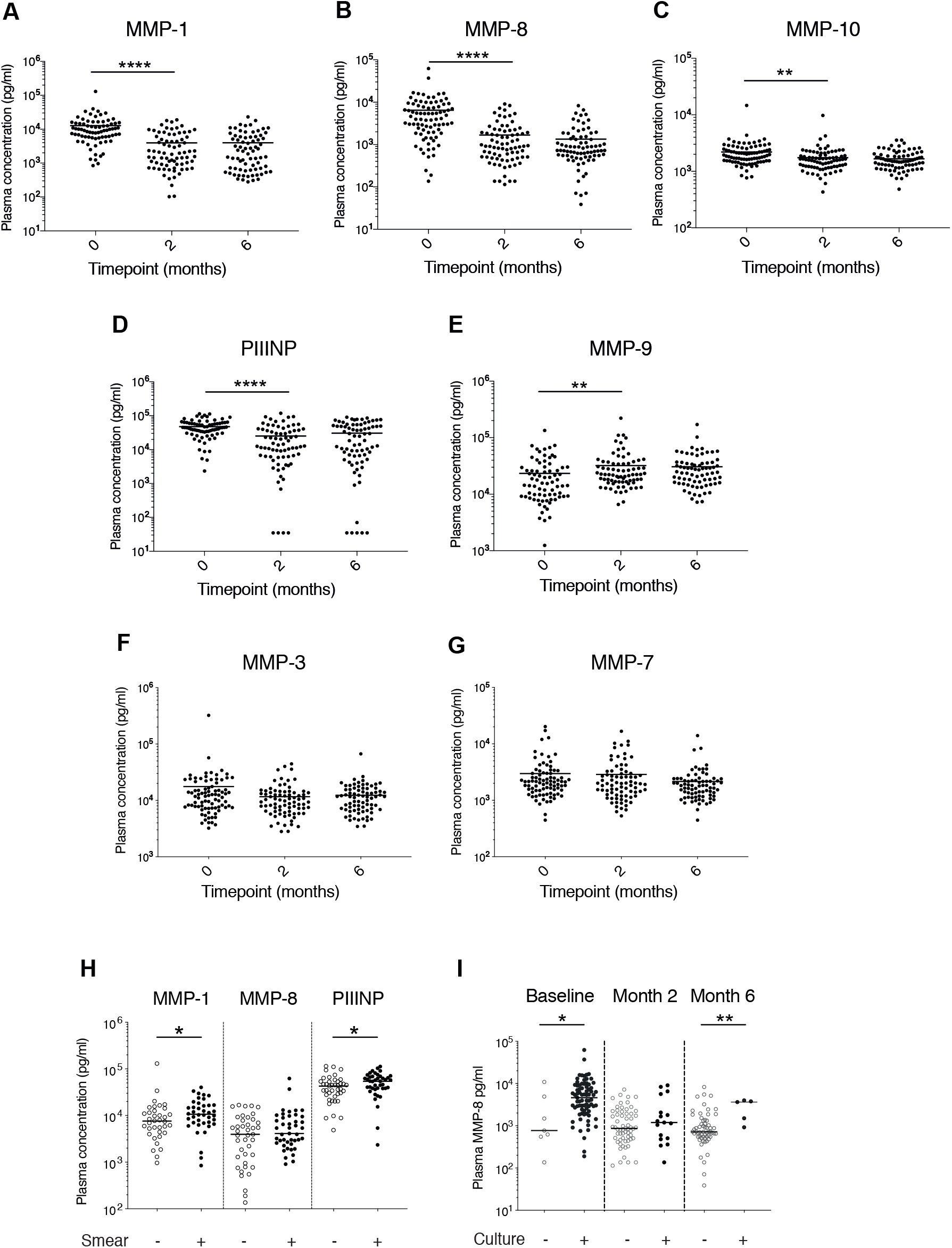
Plasma MMP & PIIINP concentrations during TB treatment. A-D: Plasma MMP-1, -8, -10 and PIIINP concentrations decreased between baseline (TB diagnosis) and the end of month 2 of TB treatment but did not decrease further between month 2 and month 6. E: Conversely, plasma MMP-9 increased between baseline and month 2. F-G: Plasma MMP-3 and MMP-7 did not change over time H: Plasma MMP-1 and PIIINP, but not plasma MMP-8, were increased in participants who were smear positive compared to smear negative at TB diagnosis. I: Plasma MMP-8 was increased in *Mycobacterium tuberculosis* sputum culture positive compared to culture negative participants at TB diagnosis and at the end of month 6 post TB treatment initiation. Analysis was by Kruskal-Wallis test with Dunn’s Multiple Test comparison. P values are summarised: * p<0.05, ** p<0.001, **** p<0.0001. Where no p value is reported, p>0.05.

### Elevated plasma MMP-1 & PIIINP in smear positive disease

Plasma MMPs and PIIINP were compared in sputum smear positive participants and sputum smear negative participants at baseline (Figure panel H). Plasma MMP-1 and PIIINP were significantly increased in sputum smear positive compared to sputum smear negative participants. Plasma MMP-3, -8, -9 and -10 did not differ by smear status. A longitudinal analysis by smear status was not performed as only one participant had a subsequent smear positive result during treatment.

### Plasma MMP-8 associates with sputum culture status at TB diagnosis and month 6

At the end of month 6, five (5.8%) participants remained sputum culture positive for *Mtb*, on liquid culture. Four out of five of these participants were HIV negative. *Mtb* culture confirmed that in two cases drug-sensitive isolates at diagnosis remained drug-sensitive at month 6, whilst in two cases isolates that were isoniazid-resistant at TB diagnosis were also rifampicin-resistant at month 6, indicating the development of MDR TB. In one case, drug susceptibility test results were available neither at diagnosis, nor later timepoints. Only one of these patients had a positive result on smear microscopy at month 6, and all cases were smear negative at month 2, indicating that the majority of these cases would not have been identified by current microscopy-based methods of screening for treatment failure.

Plasma MMPs were compared in sputum culture positive and culture negative participants at each timepoint (Figure panel I and Supplementary Figure S1). Plasma MMP-8 was significantly increased in *Mtb* culture positive compared to culture negative participants at TB diagnosis (median 4609 pg/ml, IQR 2353-9048 vs median 775 pg/ml, IQR 551-4920, p=0.019 respectively) and also at month 6 (median 3650 pg/ml, IQR 1214-3888 vs median 720 pg/ml, IQR 551-1321, p=0.008). However, there was no significant difference found at month 2 (median 1194 pg/ml, IQR 493-3846 for sputum culture positive vs 870 pg/ml, IQR 499-1986 for sputum culture negative, p=0.379). No other MMP, nor PIIINP concentration, differed by sputum culture status at any of the three timepoints. By ROC curve analysis, plasma MMP-8 at month 6 predicted month 6 sputum culture status with an area under the curve of 0.847, corresponding to a sensitivity of 100% and a specificity of 65% at the optimal cut-off (>920 pg/ml) (Supplementary Figure S2).

## Discussion

In this longitudinal analysis of TB patients on treatment, we found that plasma MMP-1, -8, -10 and PIIINP decreased with effective TB treatment over a period of two months. Whilst all but one participant in this study converted to smear negative by the end of six months of TB treatment, five participants were culture positive at six months. Notably, only one of these participants would have been identified by standard sputum smear microscopy-based screening at six months. Elevated plasma MMP-8 at TB diagnosis and at the end of six months TB treatment was associated with sputum culture positivity, indicating that plasma MMP-8 is a candidate biomarker for monitoring treatment response.

Neutrophils are a potential source of MMP-8, which may be stored in granules before release. *In vitro*, neutrophils secrete MMP-8 directly in response to *Mtb* infection in a dose-dependent manner, and in response to cellular networks (13). We have previously demonstrated that elevated plasma MMP-8 is associated with lipoarabinomannan positivity and neutrophil count in HIV-associated TB (8). In patients starting TB treatment and then antiretroviral therapy for HIV who go on to develop paradoxical TB-IRIS, plasma MMP-8 is also increased at TB diagnosis and at TB-IRIS presentation (8). Together, these findings suggest that plasma MMP-8 may be a surrogate plasma marker of mycobacterial load and neutrophil-driven immune responses in TB.

This study highlights the problem of identifying treatment failure in TB. Despite being started on TB treatment, five patients in the study remained culture positive for *Mtb* at six months. The majority of these would not have been identified by standard smear-based methods of assessing for treatment failure. A plasma biomarker, such as MMP-8, could provide a useful additional objective risk indicator to alert treating clinicians to the possibility of treatment failure, especially where resources are limited and in the case of sputum non-productive patients. If further developed for measurement using a low-cost point-of-care tool, for example a lateral flow device, it could be implemented at the community level as a rule-out triage test, whereby a low reading supports treatment success, and a high reading prompts repeat culture.

This study is not the first to identify an association of MMP-8 with culture positivity in TB patients on treatment. A study by Sigal *et al*. identified an association of elevated ratios of serum MMP-8 at week 8 to baseline MMP-8 with culture positivity at week 8 and week 12 but did not examine later timepoints (14). Another study by Lee *et al*. evaluated a number of potential biomarkers in plasma at baseline and two months (15). At month 2, MMP-8 concentrations were increased in patients who were culture positive compared to culture negative, with an AUC of 0.632 on ROC curve analysis. This association is consistent with our findings, but at a different time point. An explanation for this difference is that Lee *et al*. included only participants with drug-sensitive TB, without HIV infection. We have previously reported elevated plasma MMP-8 associated with paradoxical TB-IRIS, which occurs in the first months of TB treatment in people living with HIV infection who start ART after TB treatment initiation (8). We did not evaluate the occurrence of TB-IRIS in this study.

A strength of this cohort study was the detailed microbiological follow up and inclusion of both HIV seropositive and seronegative participants, as well as drug-susceptible and drug-resistant TB cases. However, this was an exploratory study as opposed to a diagnostic accuracy assessment, and further evaluation is required to discern the clinical utility of these findings. It is also important to recognise that additional clinical factors, including symptoms and BMI monitoring may indicate patients who are failing TB treatment. We did not evaluate these indicators in this cohort.

In conclusion, we describe an association of plasma MMP-8 with sputum *Mtb* culture positivity at the beginning and end of TB treatment, in a cohort of patients of mixed HIV serostatus. We advocate the further evaluation of plasma MMP-8 as a biomarker of culture positivity to support TB treatment monitoring as a triage test, with the aim of early identification of treatment failure and appropriate allocation of diagnostic resources, to better support care of TB patients and improve treatment outcomes.

## Data Availability

All data produced in the present study are available upon reasonable request to the authors.

## Acknowledgements

Funding was from the British Infection Association; National Institute for Health Research UK Starter Grant for Clinical Lecturers (Academy of Medical Sciences UK, Wellcome, Medical Research Council UK MR/P023754/1, British Heart Foundation, Arthritis Research UK, Royal College of Physicians, and Diabetes UK); National Institutes of Health (U01 AI 069924, K08 AI106420); the US Civilian Research and Development Foundation (OISE-16-62061-1); and the South African Medical Research Council (grant number #RFA –CC: TB/HIV/AIDS-01-2014). We would like to thank Fernanda Maruri for assistance with study data collection.

