## Supplementary Table and Figures for "Elevated Plasma Matrix Metalloproteinase-8 associates with Sputum Culture Positivity in Pulmonary Tuberculosis"

#### Supplementary Table S1 Spearman r correlation between analytes

| | MMP-1 | | | MMP-3 | | | MMP-7 | | | MMP-8 | | | MMP-9 | | | MMP-10 | | |
|---|---|---|---|---|---|---|---|---|---|---|---|---|---|---|---|---|---|---|
| | r | 95% CI | p value | r | 95% CI | p value | r | 95% CI | p value | r | 95% CI | p value | r | 95% CI | p value | r | 95% CI | p value |
| MMP-3 | 0.125 | -0.009 to 0.255 | 0.059 | | | | | | | | | | | | | | | |
| MMP-7 | **0.244** | 0.114 to 0.366 | **0.000** | 0.024 | -0.106 to 0.154 | 0.709 | | | | | | | | | | | | |
| MMP-8 | **0.377** | 0.256 to 0.486 | **0.000** | 0.097 | -0.033 to 0.225 | 0.132 | 0.172 | 0.043 to 0.296 | 0.007 | | | | | | | | | |
| MMP-9 | **-0.385** | -0.494 to -0.265 | **0.000** | -0.031 | -0.164 to 0.103 | 0.642 | -0.099 | -0.230 to 0.0352 | 0.136 | 0.197 | 0.065 to 0.323 | 0.003 | | | | | | |
| MMP-10 | 0.165 | 0.032 to 0.293 | 0.013 | **0.567** | 0.471 to 0.649 | **0.000** | 0.160 | 0.030 to 0.284 | 0.013 | **0.197** | 0.069 to 0.319 | **0.002** | -0.059 | -0.191 to 0.076 | 0.379 | | | |
| PIIINP | **0.759** | 0.696 to 0.811 | **0.000** | 0.086 | -0.045 to 0.215 | 0.184 | 0.158 | 0.028 to 0.283 | 0.015 | **0.224** | 0.096 to 0.345 | **0.000** | **-0.407** | -0.513 to -0.289 | **0.000** | 0.069 | -0.063 to 0.198 | 0.290 |

Abbreviations: Confidence interval (CI); Matrix metalloproteinase (MMP); Procollagen III N-terminal propeptide (PIIINP). Data were included from all timepoints.

To correct for multiple comparisons a p value <0.00238 was considered significant and these results are highlighted in bold.

### Supplementary Figures

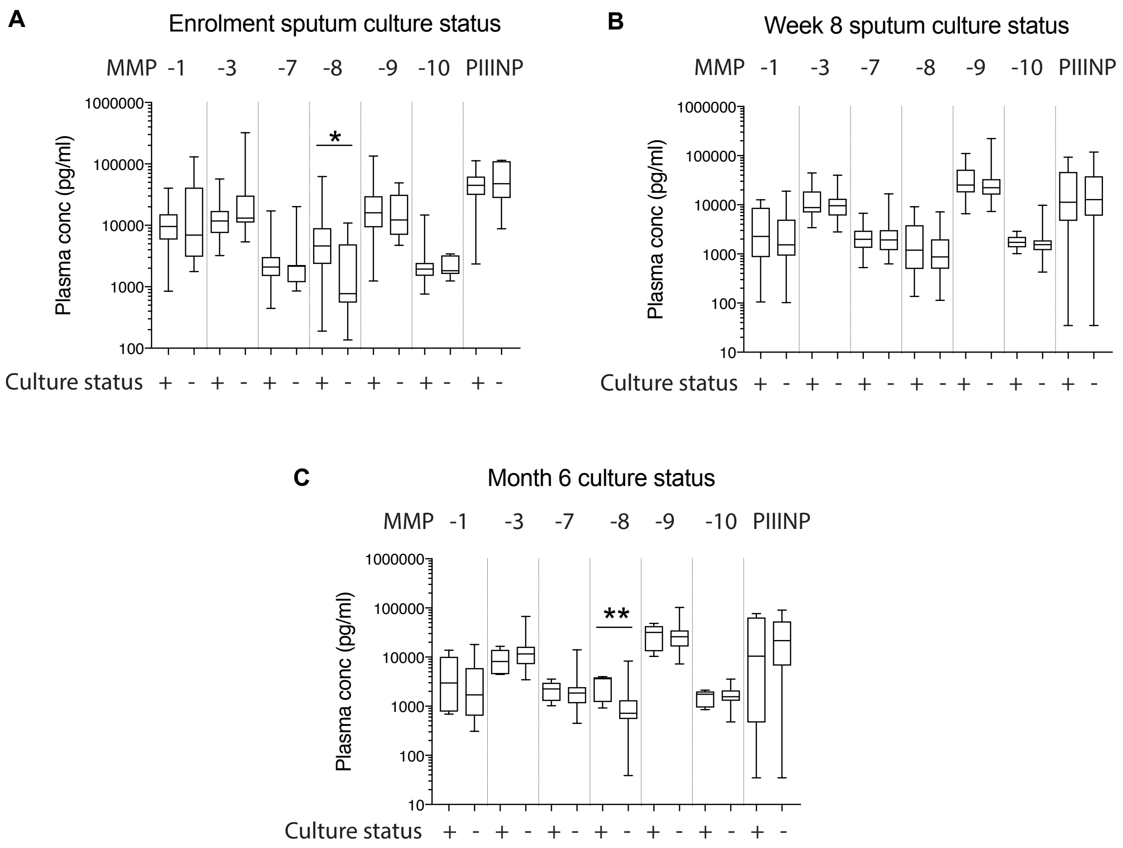


Supplementary Figure S1 Plasma MMPs and PIIINP by sputum *Mycobacterium tuberculosis* culture status

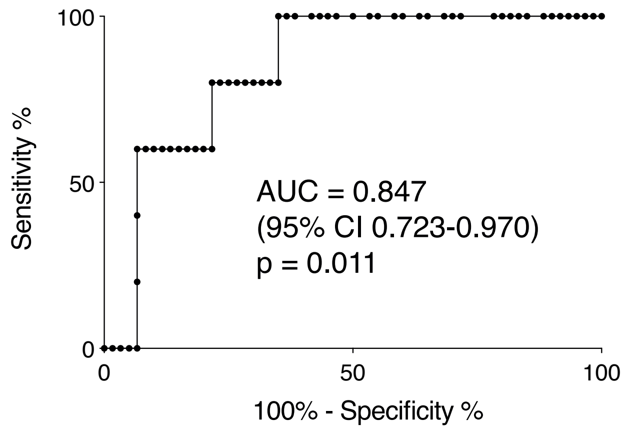


Supplementary Figure S2 Receiver operating characteristic (ROC) curve analysis of MMP-8 concentration at month 6 post-TB treatment initiation for identification of culture positivity
